# A new method to triage colorectal cancer referrals using serum Raman spectroscopy and machine learning

**DOI:** 10.1101/2020.05.20.20108209

**Authors:** Cerys A Jenkins, Susan Chandler, Rhys A Jenkins, Kym Thorne, Freya Woods, Andrew Cunningham, Kayleigh Nelson, Rachel Still, Jenna Walters, Non Gwynn, Wilson Chea, Rachel Harford, Claire O’Neill, Julie Hepburn, Ian Hill, Heather Wilkes, Greg Fegan, Peter R Dunstan, Dean A Harris

## Abstract

Suspected colorectal cancer (CRC) referrals based on non-specific symptoms currently lead to large numbers of patients being referred for invasive investigations and poor yield in cancer detection. Secondary care diagnostics, particularly endoscopy, struggle to meet the ever-increasing demand and patients face lengthy waits from the point of referral. Here we propose a blood test utilising high-throughput Raman spectroscopy and machine learning as an accurate triage tool. We present results from the first mixed methods clinical validation study of its kind, evaluating the ability of the test to perform in its target population of primary care patients, and its acceptability to those administering and receiving the test. The test was able to accurately rule out cancer with a negative predictive value of 98.0%. This performance could reduce the number of invasive diagnostic procedures in the cohort by at least 47%. Collectively, our findings promote a novel, non-invasive solution to triage CRC referrals with potential to reduce patient anxiety, accelerate access to treatment and improve outcomes.

## Introduction

Colorectal cancer (CRC) is the second largest cause of cancer related death worldwide.^1^ The majority (54%) of cases of CRC in the UK are diagnosed through patients presenting to primary care with bowel symptoms.^2^ Patients have to satisfy strict referral criteria in line with NICE guidance to be referred along the ‘Urgent Suspected Cancer’ (USC) pathway.^3,4^ The USC pathway was introduced to standardise referrals and investigations for suspected cancer to reduce time to diagnosis. The pathway recommends that patients see a specialist within two weeks and receive their first treatment for cancer within 62 days.

However, the symptoms for CRC are non-specific and are shared by a number of benign conditions such as irritable bowel syndrome and haemorrhoids. The USC pathway is based on a positive predictive value (PPV) for cancer of only 3%. The lack of specificity leads to large numbers of patients being referred along the pathway and needing investigation. Increasing referral rates have contributed to demand for colonoscopy (the gold standard diagnostic test for CRC) doubling over the last five years with numbers set to increase further.^5^

Prompt recognition and reporting of symptoms by the public has been advocated for CRC, but has only succeeded in further increasing referral rates for investigation, with no impact on rates of earlier disease diagnosis and little overall reduction in CRC mortality.^5,6^ The current USC pathway has failed to have a significant effect on the ability to detect CRC earlier and change the outcomes of CRC.^7,8^ A blood test combining high-throughput serum Raman spectroscopy (RS) and machine learning analysis could transform the CRC referral pathway. Our development of a blood test to improve triage of referrals on the USC pathway is timely as shown by the top three research priorities of The Detecting Cancer Early Priority Setting Partnership namely, (1) what simple, non-invasive, painless, cost-effective, and convenient tests can be used to detect cancer early? (2) can a blood test be used to detect some or all cancers early, and how can it be included into routine care? and (3) would increasing access to tests to diagnose cancer within General Practices improve the number of cancers detected early, and is it cost effective?^9^

RS is a vibrational spectroscopic technique that provides rapid, cost-effective analysis of biological samples. RS simultaneously measures a range of molecular species (proteins, nucleic acids, lipids, etc) within biological samples to produce a spectrum or ‘biochemical fingerprint’ unique to the sample (Figure 1). The spectrum can be considered a snapshot of a patient’s health or disease status at a given time. Spectral data can be coupled to machine learning to develop discriminatory or identification models^10,11^. Applications of RS for the detection of diseases in tissue and biofluid samples have previously been reported for a wide range of diagnostic applications including breast cancer, ^12^ brain tumours,^13^ bladder cancer,^14^ oral cancer^15^ and CRC^16^. Whilst the results are promising, approaches differ and studies to date have been largely limited to pilot studies.^17,18^

**Figure 1:**
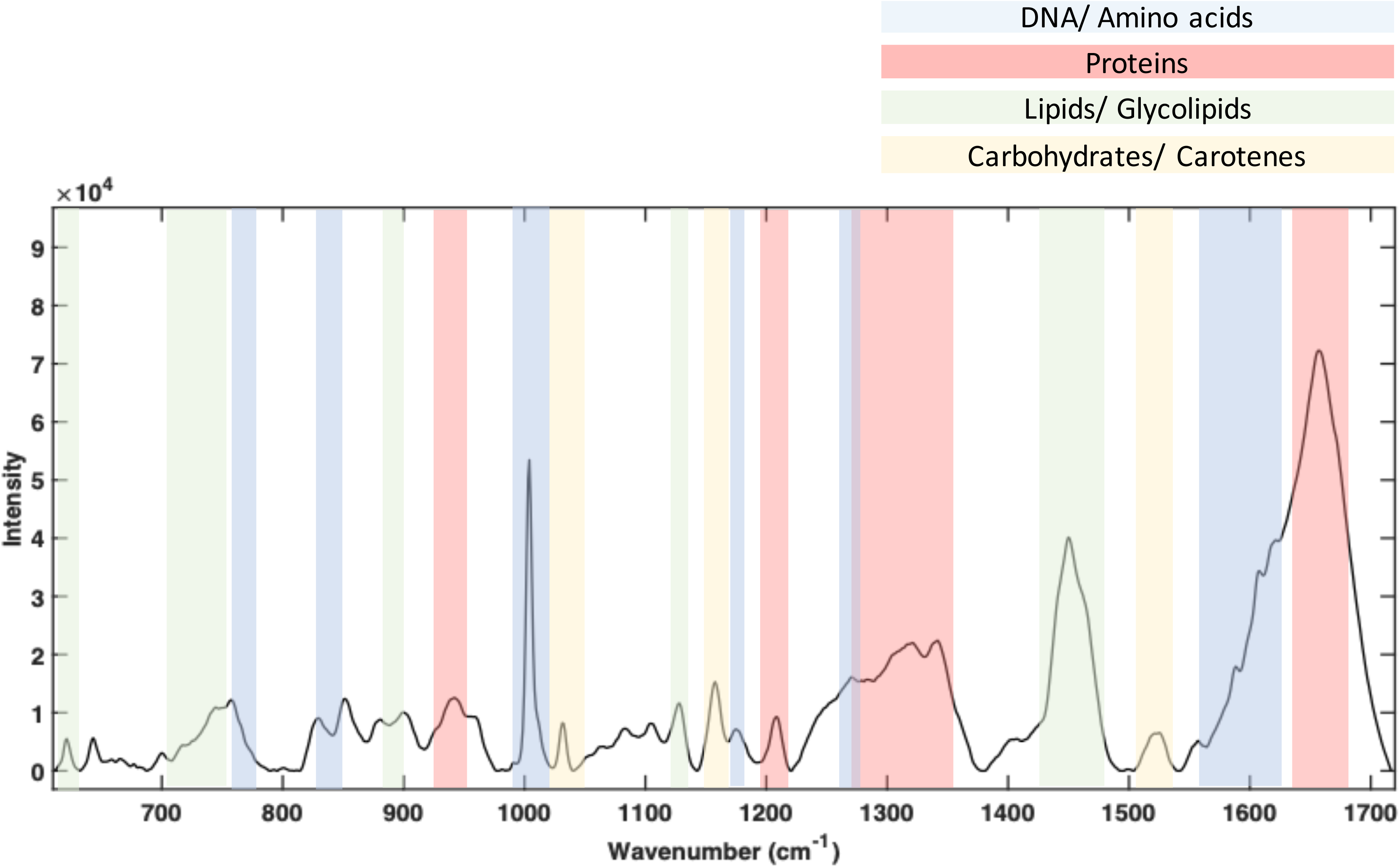
A typical liquid serum Raman spectrum collected with 785 nm laser excitation. Spectrum was collected using Renishaw Wire^TM^ (version 4.1) from 200 *μl* of serum using high-throughput substrate platform.^16^ Equipment: Renishaw In Via Raman spectrometer.

Our application to CRC has developed a high-throughput (HT) Raman spectral measurement platform that can be applied to liquid serum samples.^16^ The ability to measure samples in a liquid state holds an advantage over other vibrational spectroscopic methods such as Fourier transform infrared (FTIR) spectroscopy studies where serum samples must be dried prior to analysis.^19^ This allows Raman spectral measurements to be collected and analysed in a rapid timeframe without the need to wait for sample preparation.

The HT Raman platform was used to collect data from a retrospective cohort of 100 patients with known clinical outcomes of CRC or non-cancer (control). The data from these patients (training set) were used to develop the machine learning based Raman-CRC model.

For a disruptive technology such as Raman-CRC to translate to the proposed clinical setting it is crucial that it gains acceptance with patients and end-users (General Practitioners; GPs). To ensure patient relevance two former patients from the Public Involvement Community (JH and IH) were involved in the design of the study to ensure the study was relevant to patient needs. Clinical attitudes towards the test were explored via a qualitative evaluation of attitudes of the service end-users (GPs), exploring the use of Raman-CRC as a triage tool and the potential of Raman-CRC fitting into multiple areas of the clinical pathway.

Here, we present preliminary results of the first application of the Raman-CRC model to Raman spectra of serum samples from the largest prospective cohort to date (n=535). This study presents results from the first mixed methods approach for this indication including a nested qualitative study (Figure 2). It considers both the utility of a Raman-CRC blood test to streamline the referral pathway for suspected cancer patients and explores its potential to translate into a clinical setting through end user focus groups.

**Figure 2:**
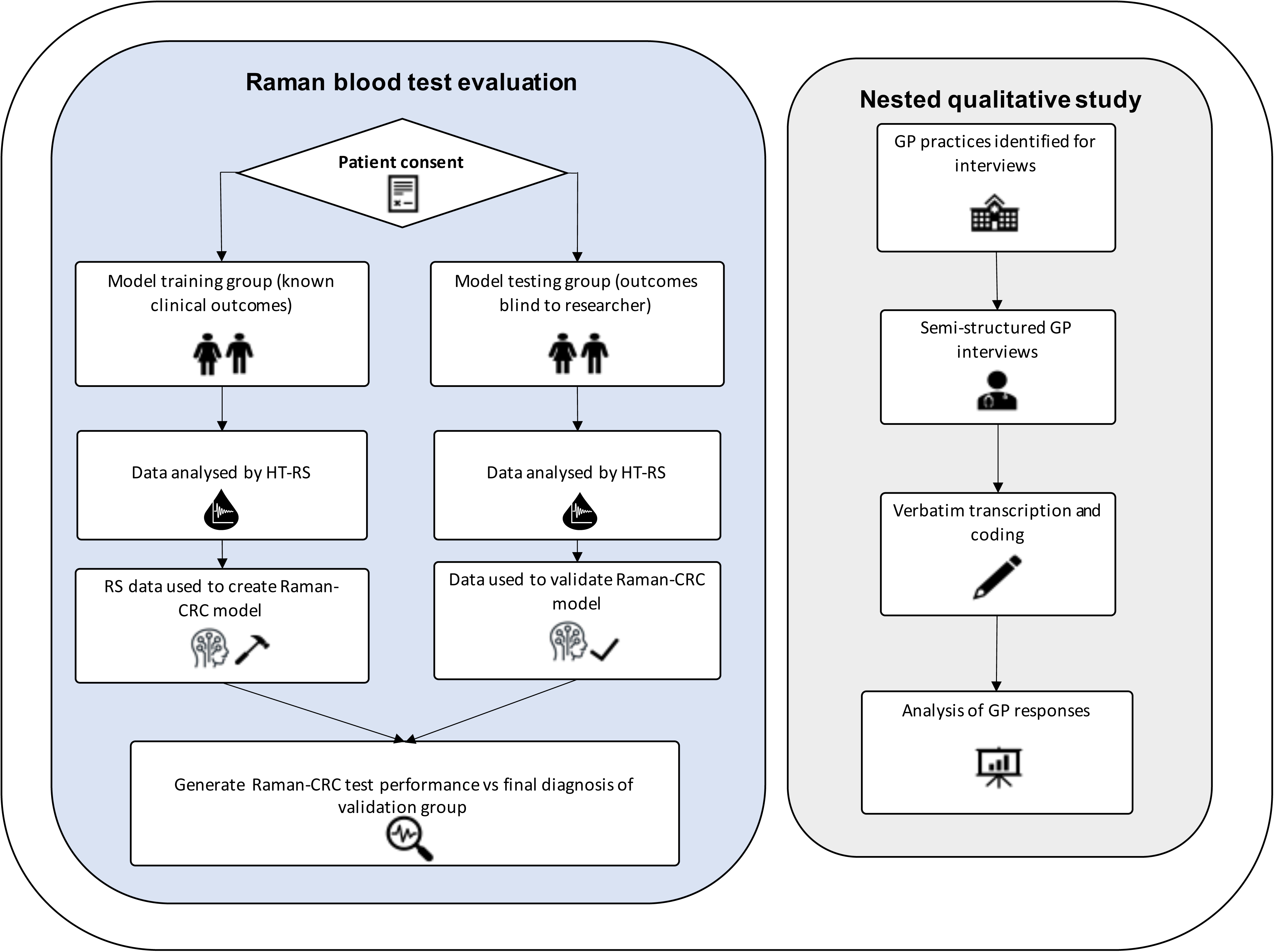
This mixed methods prospective clinical validation study incorporated a retrospective cohortanalysis to build the Raman-CRC model, the prospective study for clinical validation and a nested qualitative study for investigating attitudes of the test in primary care.

## Results

### Development of the Raman-CRC diagnostic model with retrospective patient cohort

Data from a previously recruited retrospective cohort of 100 patients with known clinical diagnosis (CRC or control) was used to develop the Raman-CRC blood test model. Patients were age and sex matched where possible (Table 1). Serum samples from the cohort were analysed using a HT Raman platform to provide 5 biological repeat spectra from each patient. In total this provided 500 spectra (250 CRC; 250 control) for analysis. When coupled to machine learning this data was used to train classification models to detect the spectral differences between CRC and control blood sera and then predict the disease status of unknown samples. The results from the training and testing (blind test set) of a random forest (RF) classification are presented (Figure 3). The accuracy and receiver operator characteristic (ROC) curve for the diagnostic model was calculated from a 20% blind holdout validation patient set. The holdout set were excluded from model building and tested against the random forest algorithm which was trained with the remaining 80% (whereby patients can only appear in either the training or cross validation set). The Raman-CRC model showed a good area under the curve (AUC) of 0·84 and performed with a sensitivity and specificity of 84.0% and 78·0% respectively, indicating the ability to distinguish patients with and without CRC within the prospective cohort (Figure 3).

**Figure 3:**
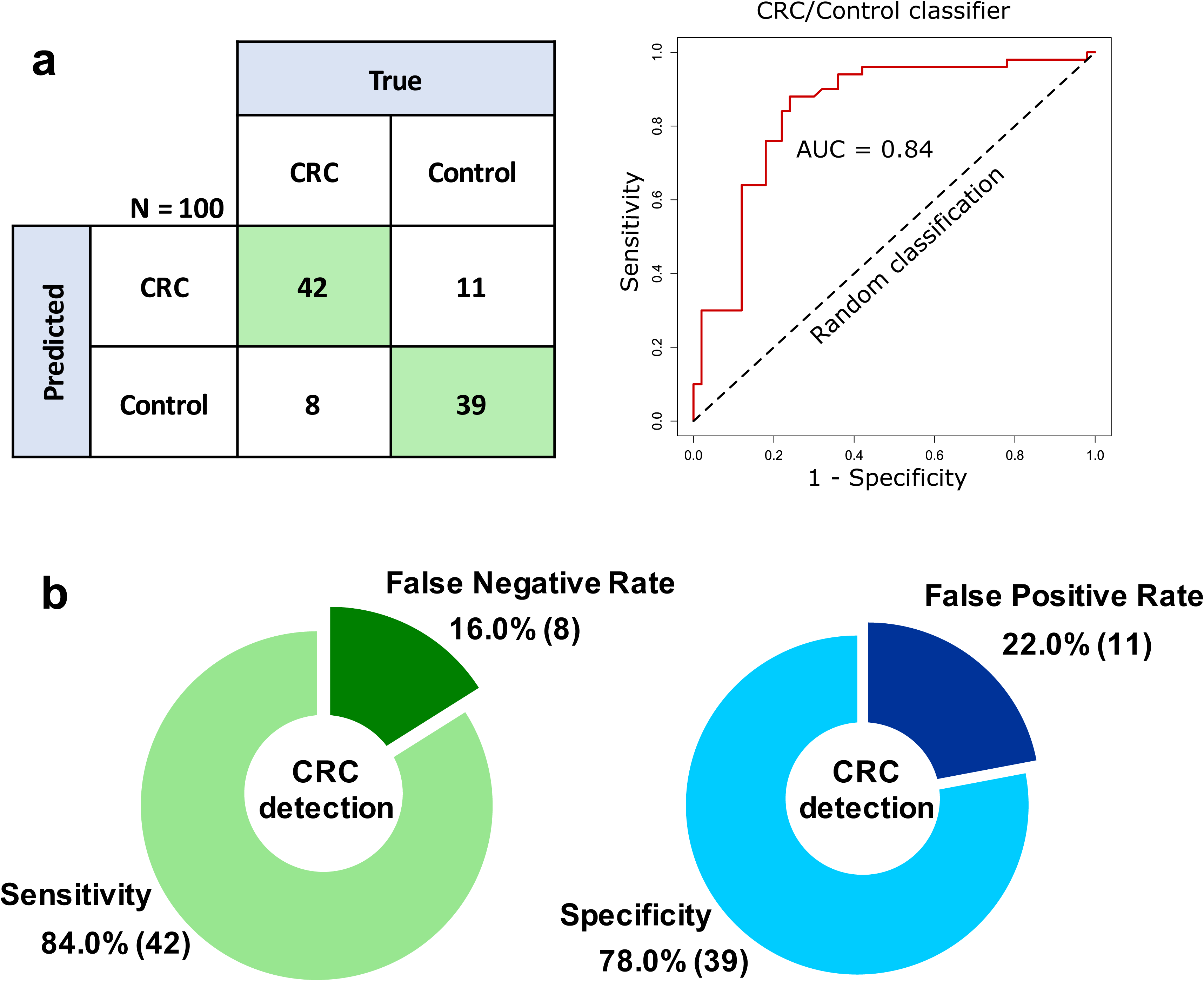
Raman-CRC random forest model training and performance analysis. **a** confusion matrix and ROC curve analysis of the model training CRC and control. All data refer to resampled and averaged test set predictions and **b** sensitivity, specificity, false negative rate and false positive rate for the Raman-CRC model.

**Table 1:** Retrospective cohort information

|  | Colorectal cancer | Control |
| --- | --- | --- |
| Total patients | 50 | 50 |
| Mean Age (years), (SD) | 67 (11) | 64 (13) |
| Female | 25 | 26 |
| Male | 25 | 24 |

Control patients were confirmed not to have CRC following diagnostic colonoscopy. Sex, age and final diagnosis of the patients were matched wherever possible.

### Prospective validation study

Following promising results from the Raman-CRC model development, analytic researchers next tested the prospective USC GP patient samples against the trained model to perform analysis representative of the target end-use of Raman-CRC.

The study captured a wide variance of cases within the total 535 patients from primary care including patients with non-cancer diseases, pre-cancerous polyps and other cancer types (Table 2). In accordance with prevalence in a GP population 29 patients (5%) were diagnosed with CRC through the traditional referral pathway. Patient ages were comparable between the CRC and the non-CRC group. A predominance of male patients was observed in the CRC group consistent with its known distribution. Data capture from the prospective cohort allowed a comparison of the symptomatic presentation of the patients (Table 3). Minimal difference in symptom frequency or routine blood results (haemoglobin, ferritin) between cancers and non-cancers was observed highlighting the lack of clinical specificity.

**Table 2:** Prospective primary care cohort with final diagnosis breakdown

|  | Tumour location | Initial diagnostic test |  | Total |
| --- | --- | --- | --- | --- |
|  |  | Colonoscopy | CT colonogram |  |
| Colorectal tumours |  |  |  |  |
|  | Caecal/<br>ascending colon | 6 | 2 | 8 |
|  | Sigmoid | 7 | 1 | 8 |
|  | Rectal | 10 | 3 | 13 |
| Other tumours |  |  |  |  |
|  | Pancreatic | 1 | 3 | 4 |
|  | Prostate | 2 | 1 | 3 |
|  | Lung | 1 | 2 | 3 |
|  | Bladder | 1 | 1 | 2 |
|  | Renal | 0 | 1 | 1 |
|  | Peritoneal/ovarian | 0 | 1 | 1 |
|  | Breast | 1 | 1 | 2 |
|  | Heptocellular | 0 | 1 | 1 |
|  | NET | 0 | 1 | 1 |
|  | Anal SCC | 1 | 1 | 2 |
| Non-malignant disease |  |  |  |  |
|  | Colorectal polyps | 90 | 12 | 102 |
|  | Colitis | 4 | 0 | 4 |
|  | Ovarian Cyst | 1 | 0 | 1 |
| Control |  | 228 | 151 | 379 |
|  |  |  | **Total** | **535** |

**Table 3:** Prospective cohort information with presenting symptoms

|  |  | CRC diagnosis | Non-CRC diagnosis |
| --- | --- | --- | --- |
|  | Participants, n | 29 | 506 |
|  | Male | 23 | 241 |
|  | Female | 6 | 265 |
|  | Median age (range) | 71 (51-87) | 70 (50-92) |
| Presenting symptom: |  |  |  |
|  | rectal bleeding | 15 (52) | 163 (32) |
|  | change in bowel habit | 19 (66) | 414 (82) |
|  | loose stool | 11 (36) | 282 (56) |
|  | increased frequency | 13 (45) | 217 (43) |
|  | urgency | 4 (13) | 82 (16) |
|  | incomplete emptying | 4 (14) | 84 (17) |
|  | constipation | 6 (20) | 162 (32) |
|  | abdominal pain | 9 (30) | 195 (39) |
|  | anal pain | 2 (7) | 23 (5) |
|  | abdominal mass | 1 (3) | 21 (4) |
|  | rectal mass | 3 (10) | 16 (3) |
|  | anal mass | 1 (3) | 12 (2) |
|  | loss of appetite | 3 (10) | 59 (12) |
|  | weight loss | 10 (33) | 152 (30) |
| Blood test markers |  |  |  |
|  | Haemoglobin (median;range) | 125(71-161) | 133 (63-207) |
|  | Ferritin (median;range) | 30 (6-617) | 75 (4-2427) |
|  | CEA (median;range) | 6 (1-2385) | 2 (1-1149) |

Values are n(%) unless otherwise stated.

After patient exclusions (detailed in supplementary information Figure 1), 408 patients remained whose CRC or non-CRC diagnosis was based upon an initial investigation of colonoscopy or CT colonography. Raman spectra from these patient samples were collected via the HT data collection platform. Data were then analysed, with the researcher blinded to patient diagnosis, using the Raman-CRC model on a spectrum by spectrum basis. This produced a probability for each spectrum of being cancer which was converted to a patient-wise result by averaging the results to produce an overall probability of a patient having cancer. The output from the model prediction for each patient was then compared to their final diagnosis to produce performance metrics for the test when compared by patient initial investigation (Figure 4).

**Figure 4:**
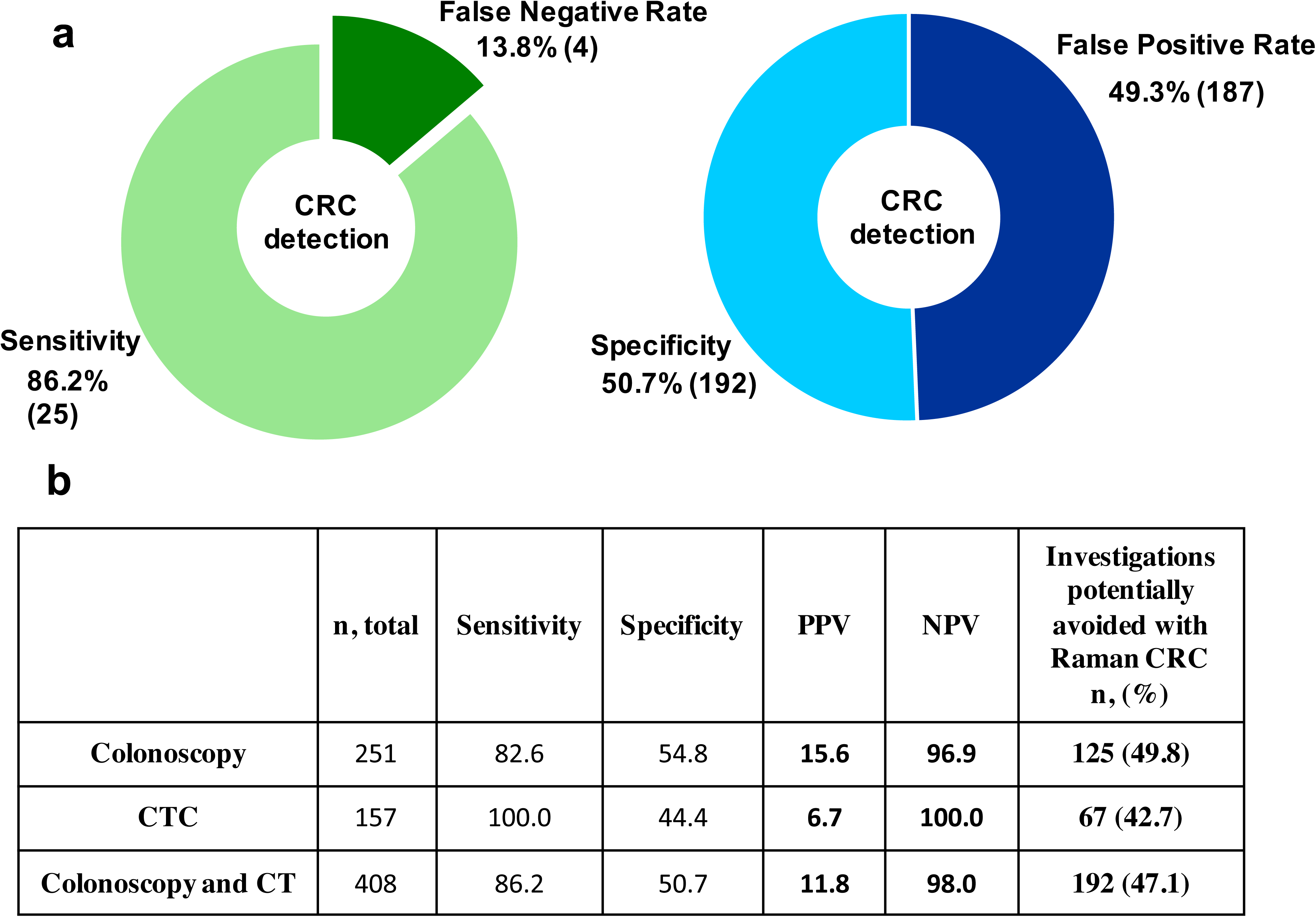
Disease prediction for the prospective validation cohortfrom secondary care USC referral patients, **a** overall sensitivity, specificity, false negative rate and false positive rate for the Raman-CRC model on a per-patient basis following blind analysis and **b** breakdown of the model performance according to initial diagnostic test, including NPV and PPV values calculated from confusion matrices (Supplementary tables S.2-4) and number of investigations that could have been avoided if Raman-CRC had been used to triage the referrals.

The negative predictive value (NPV) for the cohort of 98% can be considered excellent and within a GP setting gives the power to ‘rule out’ CRC. The PPV figure of 11.8% is a marked improvement on the current symptomatic pathway that has a PPV of just 3%.^3^

Prospective recruitment and detailed patient data capture has contributed to a better understanding of the variability of illnesses within the target population variance for the Raman-CRC test and will now inform future model development that encapsulates patients from different groups including pre-cancerous polyps. The patient comorbidities and tumour site in relation to the Raman-CRC result was also investigated in the colonoscopy cohort. There was no clear correlation between patient comorbidities and medication against the performance of the Raman result. Raman-CRC performed well across all colonic areas, with a pickup rate for cancer of 100·0% from the caecum to the sigmoid colon. It was less accurate at detecting rectal cancers with a 69·2% pick up rate (Supplementary table S.5).

To explore the potential economic benefit of using Raman-CRC as a triage tool for secondary care testing a preliminary cost analysis was conducted. With reference to patients investigated initially by colonoscopy, if the Raman-CRC test had been used, 49.8% of investigations could have been avoided on the USC pathway. In England, UK the cost of a diagnostic colonoscopy is ~£485.^20^ Initial cost analysis has estimated Raman-CRC at a cost of £40 per test and if it was performed on all patients in the cohort referred on the USC pathway receiving colonoscopy (n=251), Raman-CRC could have a direct cost saving of £50,585. A full health economic assessment is planned to evaluate future cost-effectiveness for the Raman-CRC test as a triage tool for symptomatic patients in primary care.

### Acceptability of a Raman blood test in primary care

It is important for adoption that any new diagnostic be acceptable to the end-users. To explore the acceptability of the Raman-CRC test a qualitative analysis was conducted, by focus group meetings across six primary care practices, involving 24 GPs. The mean meeting duration was 45 minutes (range 35-55 minutes) and followed a semi-structured interview format. Following analysis of the transcripts and discussion of data saturation, four key themes were identified from the discussions; perceptions of the current referral pathway, utility of Raman-CRC as a triage tool, utility of Raman-CRC as a diagnostic tool, and GP acceptability of Raman-CRC. Each key theme from the focus group discussions were then summarised (Figure 5).

**Figure 5:**
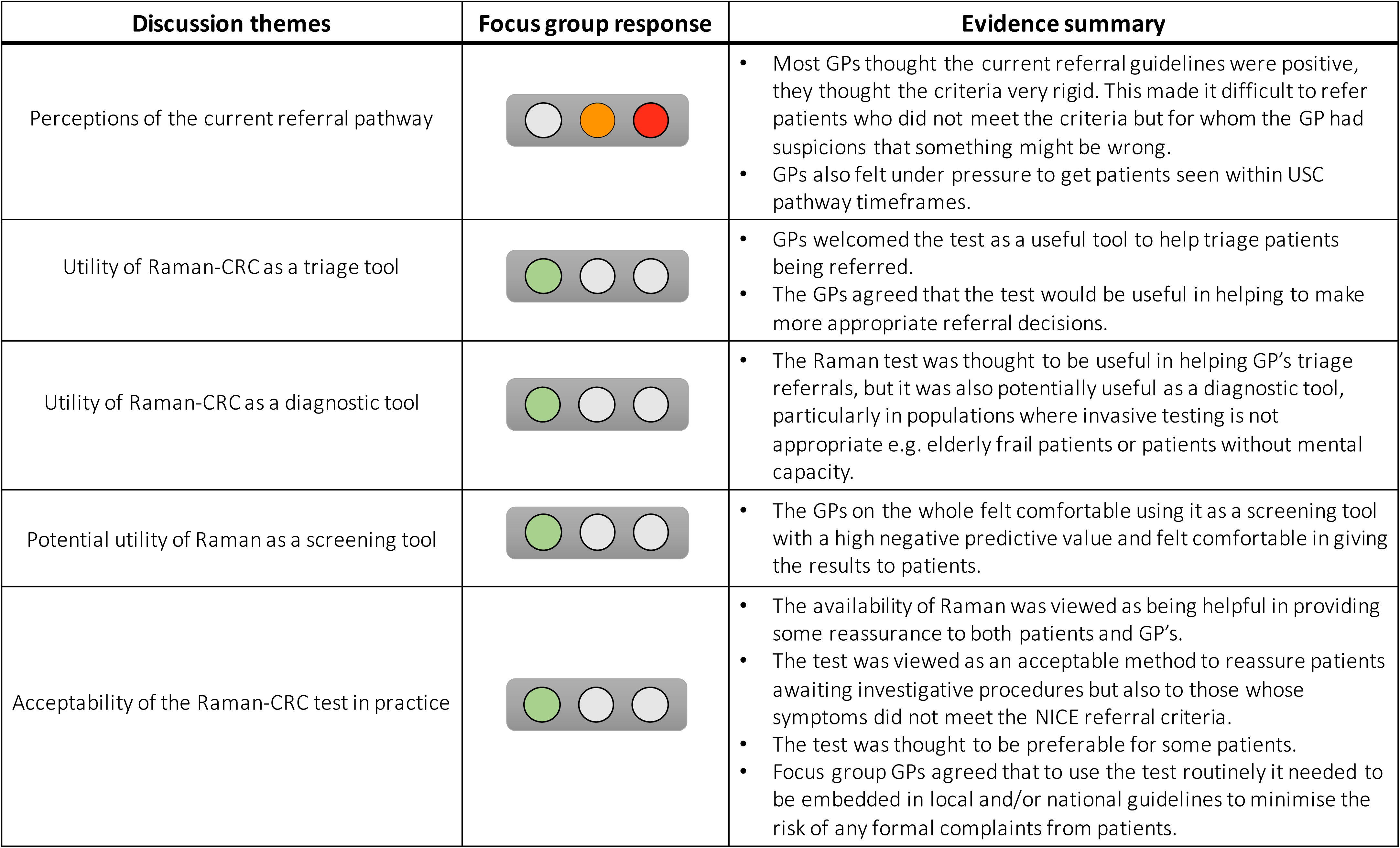
Evidence summary for primary care focus group themes.

When considering the perceptions of the current referral pathway focus group GPs agreed that they carefully considered appropriateness of USC referrals and were conscious of current capacity issues within secondary care. They highlighted patients often experiencing long waits for ‘urgent’ referrals and as such would try to “*shoehorn”* (GP 2, practice 5) patients into the USC pathway to fulfil their duty of care in a timely manner. While most GPs thought the current referral guidelines were positive, they thought the criteria very rigid. *“It doesn’t allow for atypical presentations does it? Sometimes you do just have that gut feeling when you see someone and there is no leeway to get that through.”* (GP 2, practice 4). The rigidity of the current pathway made it difficult to refer patients who did not meet the criteria but for whom the GP had clinical concerns. “I *don’t always refer to the guidelines every time because I feel that if I did I would be knocking more out than in.”* (GP 2, practice 1).

GPs welcomed the Raman-CRC test as a useful tool to help triage patients being referred and make more appropriate referral decisions. They highlighted that the test might reduce the number of unnecessary referrals. “…So *if there was a test like the Raman test and they are available it would add that reassurance./…/You may not need to do any further investigations.”* (GP 2, practice 2).

GPs also highlighted other potential uses for the test and all agreed that it would be most useful in helping to provide an evidence base for, and enabling better management of, patients who had suspect symptoms but did not meet the referral criteria. *“It’s another tool in your box. If you think it’s barn door then it doesn’t matter what a blood test shows does it?… but for those nebulous areas [it’s useful].’’* (GP 1, practice 2). It was also thought the test would go some way to helping GPs removing barriers to earlier diagnoses as evidence to refer patients *“It would be worth it to have a bit more supportive evidence if needed”* (GP 3, practice 4).

The test showed a high NPV of 98.0% showing an excellent ability to rule out cancer. It was viewed as an acceptable method to reassure patients and GPs awaiting investigative procedures and reduce anxiety in patients. *“It’s very good at saying you haven’t got cancer so you can be reassured.”* (GP 2, practice 1). The test was thought to be preferable for some patients particularly when compared to faecal based tests.

GPs also highlighted that the test has potential as a diagnostic tool in populations where invasive testing is not appropriate e.g. frail patients potentially providing a diagnosis without invasive diagnostic procedures causing harm or distress to patients. GPs on the whole felt comfortable using it as a screening tool because of the high NPV and iterated they would be comfortable providing the results to patients.

To have the confidence to use Raman-CRC routinely in primary care all agreed it needed to be adopted into local or national guidelines. Reasons for this were to minimise the risk of any formal patient complaints. “I *think it would make a lot of difference if it was in the guidance, the difficulty is at the moment is that you have got the guidance that says USC referral and [Raman] isn’t in it. /../ if it’s in the guidance you’ve got more confidence in not making a referral then.”* (GP 2, practice 2). However, GPs agreed that if the test were available and within the guidance then it would be well utilised. *“if a Raman blood test was available then I would do it, and I think you would find every GP would.”* (GP 1, practice 3).

## Discussion

We report the first prospective study to analyse blood serum with label-free RS combined to machine learning as a disruptive new technology to potentially transform the current USC pathway for CRC. The study was conducted in the intended target population for use, a symptomatic primary care population with low cancer prevalence. It shows early evidence that Raman-CRC has sufficient test performance for future utility as a symptomatic pathway triage tool in primary care. It demonstrates a good ability to exclude CRC with NPV of 98.0% and better performance than the USC pathway in predicting cancer likelihood (PPV 11.8% compared with 3%). The application of this test as a triage tool in the USC pathway showed that is has potential to reduce the number of diagnostic investigations by up to 47%. This methodology would have further benefits by reducing waiting times for diagnostic investigations, reduce patient anxiety and allow faster treatment for those more likely to have CRC.

Analysis of focus groups from primary care providers showed overwhelming support and highlighted the need for a blood test to triage primary care referrals. It gave insights into the likely clinical applications for patients missing the current criteria who they have concerns about CRC and its potential as a screening tool. GP attitudes were positive towards adoption and clinical utility for a blood-based diagnostic for CRC in primary care. The inherent reduction in patient anxiety was positively received. Test performance was considered acceptable even at this preliminary stage and would be used to influence referral behaviour if routinely available.

The Raman-CRC test accuracy is based upon a subset of the overall test cohort with the early stage binary algorithm. There were no exclusions based on comorbidities or medication which may have influenced the test performance compared with the model training data. An improved algorithm encompassing more underlying conditions and including polyps is in development and has potential for superior performance.

The International Cancer Benchmarking Partnership (ICBP) has reported the UK as having the lowest survival rates for colorectal cancer, in part through differences in diagnostic pathways and referral timelines.^21,22^ There is interest in the use of faecal immunochemical testing (FIT) in symptomatic patients in primary care with NICE guidance (DG30) supporting its use in low risk populations and pilots running in the UK for high risk groups meeting the USC referral criteria.^23^ Studies show FIT is more accurate than the NG12 USC pathway for suspected lower gastrointestinal cancer with an AUC for CRC of 0·85 compared with 0·65.^24^ Current uncertainties with widespread FIT implementation are: 1) what are the optimum cut-off levels?: 2) How acceptable a test it is to patients and GPs (with low compliance reported): 3) what its impact will be on endoscopy services, with a likely rise in demand not reduction. Unlike FIT (which detects haemoglobin) Raman-CRC has applicability to any lower GI symptom, in particular overt rectal bleeding which was the commonest presenting symptom in the ICBP study.^21^

Other emerging technologies include detection of volatile organic compounds in breath, urine and blood, and circulating tumour (ct) cells or ctDNA. Although showing promising sensitivity and specificity in known cancers, these technologies are not yet validated in target clinical populations with low cancer prevalence and are not currently cost-effective for NHS use.^25^ Raman-CRC has discernible advantages through being a rapid, reproducible, high throughput technology that is low cost (~£40 per sample) in comparison with ctDNA techniques (~£250 per sample).

A larger cohort study evaluating Raman-CRC alone and in combination with FIT (CRaFT) is underway. The CRaFT study will further develop the current diagnostic model and measure individual and combined test accuracy with FIT. It will also capture symptomatic patients’ experiences and attitudes with Raman and FIT. Future work is planned to conduct a formal cost effectiveness analysis, impact analysis in terms of earlier detection and use of downstream resources and qualitative patient and clinician test acceptability.

Raman-CRC has shown potential to become a clinician decision-making aid in symptomatic patients at higher risk for underlying CRC. It has high NPV values and shows early indications of timeframe reduction to diagnosis and resource release. The clinical impact of Raman-CRC in primary care would be twofold: 1) rapid exclusion of CRC in symptomatic patients, reducing referrals on the USC pathway, allaying anxiety and releasing colonoscopy resources; 2) to upgrade patients with low risk symptoms to the USC pathway if the Raman-CRC was positive towards earlier detection, potentially translating into improved cancer mortality. A positive test would circumvent the traditional route of outpatient review and diagnostic request by dovetailing with a ‘Straight To Test’ pathway (Figure 6).^26^. As an accurate and acceptable test Raman-CRC has the ability to transform how we detect colorectal cancer in a symptomatic population.

**Figure 6:**
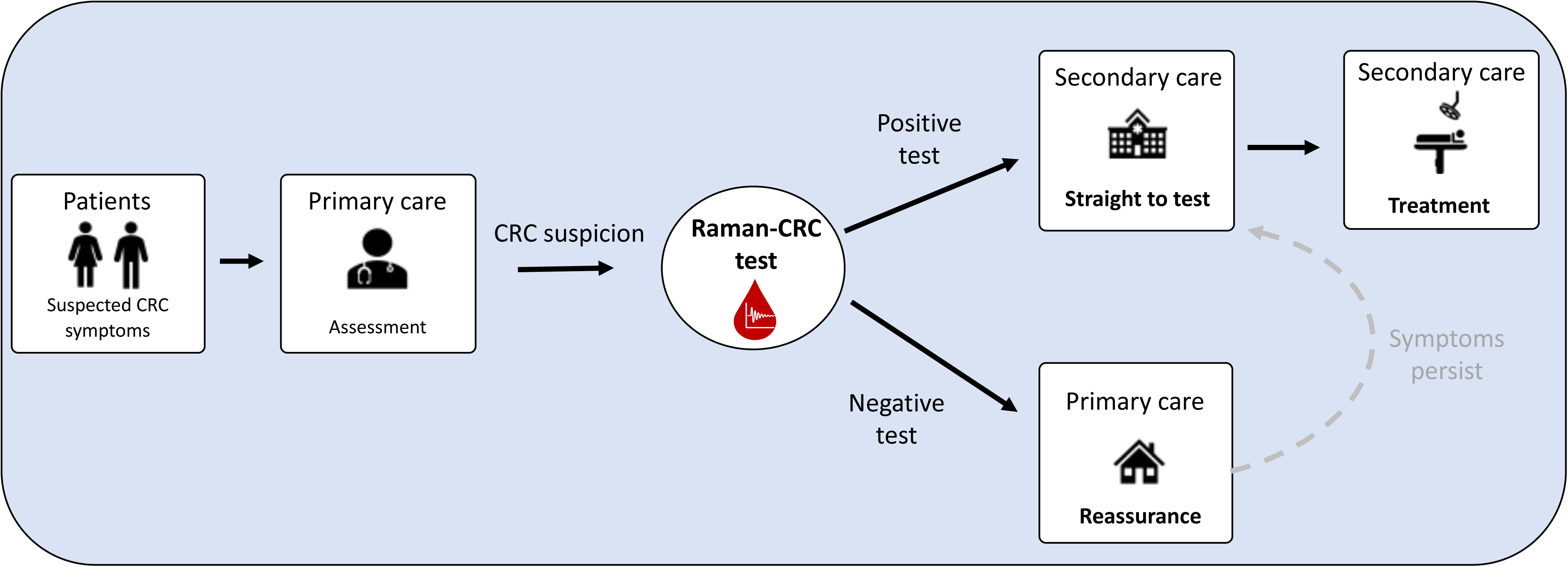
Proposed new clinical pathway incorporating Raman-CRC testing as a triage tool in primary care. Symptomatic patients with a negative Raman-CRC test are reassured in primary care, relieving pressure on secondary care diagnostic services. The pathway could lead to earlier diagnosis and a reduction in time to treatment when a positive test is combined with a straight to test pathway.

## Methods

### Study design

A prospective cohort study to evaluate the performance of Raman-CRC in primary care to triage need for referral and diagnostic testing for CRC. This work was performed as a phase 2 evaluation of clinical test performance (analytic validity in intended setting) in accordance with the CanTest framework.^27^ Results of Raman-CRC were compared to final patient diagnosis via the standard USC pathway to determine sensitivity, specificity, negative predictive value (NPV) and positive predictive value (PPV) in a symptomatic primary care population. The test results were blinded from the referring GP and patient so as to not affect the referral pathway or standard of care.

The study was conducted within Abertawe Bro Morgannwg University Health Board now Swansea Bay University Health Board (SBUHB) and managed by Swansea Trials Unit. Patient demographics, current USC pathway timelines and final diagnosis were obtained from electronic patient records (Welsh Clinical Portal) and recorded in a REDCap database.^28^ Clinical records were scrutinised up to 9 months after diagnosis to ensure missed diagnoses were captured. Results were reported according to QUADAS-2 standards.

A nested qualitative study was performed and reported according to the consolidated criteria for reporting qualitative research (COREQ) checklist involving semi-structured focus group discussions with GP practices.^29^ This explored attitudes towards the current USC pathway and the potential uses of Raman-CRC in primary care. The overall study design is summarized in Figure 2.

### Ethical approval

This study received a favourable ethical opinion by Wales REC6 (14/WA/0028). Written informed consent was obtained from all patient participants in the study and all focus group participants before interviews.

### Participants

Eligible participants were aged 50 or over and had presented to their GPs with symptoms raising suspicion of CRC as per NICE guidelines (NG12).^3^ Exclusion criteria included patients unwilling or unable to consent and patients from vulnerable groups.

### Blood sample preparation

Fasted venous blood samples were collected from patients (Vacutainer SST collection tubes BD, USA). Blood samples were centrifuged and serum aliquoted using standardised SBUHB hospital laboratory medicine workflows. Samples were aliquoted into 3 replicates and stored at −80 °C before batch analysis.

### Statistical analysis

Sample size planning (GP population - symptomatic)

The study was designed to estimate test performance of the Raman-CRC model in a population with a cancer prevalence representative of its ultimate application. A sample size of at least 75-100 patients is required as an independent blinded test set.^30^ To provide a definitive sample size for precise determination of the performance of the analysis model assuming a 10% prevalence of CRC within the cohort fulfilling USC criteria,^31^ it was estimated that the study would require 600 recruited participants based on a specificity of 81% with absolute precision of 0.1.^32,33^

### Raman spectroscopy

Serum samples were analysed using previously reported high throughput (HT) Raman methodology with modifications.^16^ Serum samples were thawed prior to analysis, liquid serum samples (200 μl) were placed into the HT platform and analysed with a 785 nm laser using a Raman microscope (InVia Renishaw, UK). All spectra were collected using the Renishaw WireTM software (version 4.1), repeat Raman spectra were obtained for each patient sample. Data collection time was between 12-15 minutes per sample.

### Raman-CRC machine learning model

Following data collection all Raman spectra underwent data pre-processing prior to further analysis. This included wavenumber calibration, data binning, smoothing, background subtraction and normalisation performed using an R^34^ package or pre-processing and was developed in-house. A random forest (RF)^35^ based machine learning model was developed using a retrospective cohort of 100 patients with known clinical outcomes of CRC or non-cancer control (Supplementary table S.1). CRC patients were confirmed to be positive for CRC by histology. Control patients in the training set were confirmed not to have CRC by colonoscopy. The RF algorithm training data used 5 repeat spectra from 50 control patients and 50 confirmed CRC patients totalling 500 spectra. The Raman-CRC model was internally cross-validated using a 20% leave-out of training data (avoiding spectra from the same patient appearing in both training and testing groups) to produce a preliminary AUC and sensitivity and specificity values.

The area under the receiver operator characteristic curve (AUC) for the model training was calculated within R from the cross validation set. The sensitivity, specificity, NPV and PPV values for Raman-CRC for the GP population were calculated within Microsoft Excel from confusion matrices comparing the final diagnosis to the Raman-CRC prediction (Supplementary tables S.2-4).

### Prospective clinical validation study

35 GP practices within SBUHB were invited to take part in the study of which 27 took part (77%). To capture patients from non-participating GP practices patients were also recruited at specialist clinics within secondary care following USC referral. Nine patients declined study participation leaving 595 patients that provided blood samples at time of consent. (Supplementary Figure 1 – STROBE diagram).

Analytic researchers were blinded to clinical information and final diagnosis for all Raman analysis of the prospective cohort data. Following pre-processing, spectra were analysed by the Raman-CRC model on a spectrum by spectrum basis. This generated a probability of CRC/control for each spectrum. The average probability for all spectra from each patient was then aggregated to produce an overall predicted probability for a patient to be positive for CRC. Any patient with a probability of greater than or equal to 0.5 was classified as CRC, and less than 0.5 non-cancer.

### Reference standard

The resultant decision for each patient produced by Raman-CRC was compared to final diagnosis as confirmed following colonoscopy or CT colonography with histological verification. Patients who did not undergo reference standard tests or had data missing from diagnostic results were excluded from analysis due to lack of reference for test performance calculations. Colonoscopy was used as the primary reference standard. The results were analysed per investigation and were separated because CT colonography is known to have a lower accuracy, with reduced ability to detect small polyps and flat cancers.^36,37^ Patients who were investigated with flexible sigmoidoscopy were excluded from analysis due to the whole colon having not been visualised.

### Primary care interviews

Semi structured focus groups were carried out at 6 primary care practices across the South Wales region (for selection criteria, demographics and identification numbers see Supplementary 2). The focus groups aimed to explore the attitudes of GPs towards current NICE guidelines, discuss the current access to diagnostic tests, level of test confidence needed in Raman-CRC before test introduction and education needs prior to test launch. Scenarios were presented during the focus groups to explore attitudes toward test application for different clinical situations with data on RS performance based on a previous pilot study in secondary care. The focus groups were conducted face to face at GP practice sites (one via video conferencing), and all GPs at each site were invited to join. GPs who participated were given the information sheet and interviews were carried out by DAH. The focus groups were audio recorded and transcribed verbatim before analysis.

### Qualitative analysis

Following checked transcription NVivo software (Version 12, QSR International Pty Ltd.) was used to code and analyse the transcripts. 3 researchers (one male, two female) independently coded the interviews to identify potential themes and the independent analyses were merged into a final coding scheme, Supplementary 2 contains the coding tree.^38^ Subthemes were generated based on consensus.

### Data availability

The datasets generated and/or analysed during the current study are available within the article and supplementary information files and available from the authors upon reasonable request.

## Acknowledgements

This study was funded through a Welsh Government Efficiency Through Technology Fund (X.481.HTT). This work was also supported through Cancer Research Wales: Raman spectroscopy and Colorectal cancer: Transforming the USC referral pathway (Registered Charitable Incorporated Organisation Number: 1167290). We would like to thank all patients for volunteering their time and donating their serum to be involved in the study.

## Clinical investigators

Department of Colorectal Surgery, Swansea Bay University Health Board: Prof J Beynon, Prof U Khot, Mr M Davies, Mr MD Evans, Mr TV Chandrasekaran, Mr GW Taylor, Mr S Ather. General practitioners (site principal investigators):

Richard Tristram, Clydach Primary Care Centre, Swansea; Heather Potter, Skewen Medical Centre, Neath; Emma Manson, Uplands Surgery, Mumbles, Swansea; David Martin Jones, Mumbles Medical Centre, Swansea; Stephen Hailey, Pennard Surgery, Swansea; Kirstie Truman, Gower Medical Practice, Swansea; Julien Bell, Grove Medical Centre, Swansea; Lynne Dowding, Kings Road Surgery, Swansea; Matthew Seager, Sketty and Killay Medical Centre, Swansea; Kirstie Truman, Mark Davies (retired), West Cross/St. Thomas’ Surgery, Swansea; Tim Evans, Fforestfach Medical Centre, Swansea; Owen Powell, Fforestfach Medical Centre (Powell practice), Swansea; Ceri Todd, High Street Surgery, Swansea; Rebecca Jones, Dulais Valley Primary Care Centre, Neath; Alistair Bennett, Dyfed Road Surgery, Neath; Steve Harrowing, Vale of Neath practice, Glynneath; Anjula Mehta, Cymmer and Cwnavon Health Centre; Richard Beynon, Llansamlet Surgery, Swansea; Richard Thomas, Kingsway Surgery, Swansea; Sherard Lemaitre, Oak Tree Surgery, Bridgend; Daniel Tacagni, Strawberry Place Surgery, Swansea; Russell Clark, Llys Meddyg, Sway Rd, Morriston; Emma Rees, Pontardawe Health Centre, Swansea; Griff Hopkin, Gowerton Medical Practice, Swansea; Duncan Williams, Amman Tawe Partnership; Alison Lilley, Castle Surgery, Neath; Mark Goodwin, Glyncorrwg Afan Valley Group Practice; Amrita Amin, St. Helen’s Medical Centre, Swansea; Dhamayanthi Vigneswaran, Cheriton Medical Centre, Swansea; Maria Cronje, North Cornelly Surgery, North Cornelly; Clare Perman, Cwmfelin Medical Centre, Neath

## Author Contributions

CAJ and SC provided study data, did the literature review, data analysis and drafted the manuscript. RJ, FW, AC, KN, WC provided study data and/or contributed to the interpretation of results. KT, DAH, PRD, JH, IH, CON, RS, NG, JW, GF have made substantial contributions to the conception and design of the work and subsequent protocol revisions; CAJ, DAH, SC, RJ, KT, FW,AC, KN, RS, JW, NG, RH, GF, HW, WC, CON, JH, IH and PRD drafted the manuscript and/or provided critical revision; approved the version submitted for publication; agree to be accountable for all aspects of the work in ensuring that questions related to the accuracy or integrity of any part of the work are appropriately investigated and resolved.

## References

1. Cancer Research UK. Bowel Cancer Statistics. https://www.cancerresearchuk.org/health-professional/cancer-statistics/statistics-by-cancer-type/bowel-cancer (accessed 03/01/20).

2. National Cancer Intelligence Network (NCIN). Routes to Diagnosis 2006–2016 workbook published version 2. http://www.ncin.org.uk/publications/routes_to_diagnosis (2016). (accessed 03/01/20)

3. National Institutue for Health and Care Excellence (Clinical guideline [NG12]). Suspected cancer: recognition and referral. NICE 2015 [updated 2017] (2017). https://www.nice.org.uk/guidance/ng12. (accessed 03/01/20)

4. Department of Health. The NHS Cancer Plan. Dep Heal. (2000); https://webarchive.nationalarchives.gov.uk/20050613200523/http://www.dh.gov.uk/PublicationsAndStatistics/Publications/PublicationsPolicyAndGuidance/PublicationsPolicyAndGuidanceArticle/fs/en?C0NTENTJD=4009609&chk=n4LXTU

5. Burki, T. K. Bowel cancer diagnostic services in the UK: at full capacity Lancet. Gastroenterol. Hepatol. 4, 15 (2019). https://doi.org/10.1016/S2468-1253(18)30393-5

6. Bowel cancer UK. Unacceptable endoscopy waiting times put launch of new world screening programme at risk https://www.bowelcanceruk.org.uk/news-and-blogs/campaigns-and-policy-blog/ending-the-capacity-crisis/. (accessed 04/02/20)

7. Mozdiak, E. et al. Systematic review with meta-analysis of over 90 000 patients. Does fast-track review diagnose colorectal cancer earlier? Aliment. Pharmacol. Ther. 348–372 (2019). https://doi.org/10.1111/apt.15378

8. Peacock, O. et al. ‘Be Clear on Cancer’: the impact of the UK National Bowel Cancer Awareness Campaign. Colorectal Dis. 8, 963–967 (2013). https://doi.org/10.1111/codi.12220

9. Badrick, E. et al. Top ten research priorities for detecting cancer early. Lancet Public Heal. (2019) https://doi:10.1016/S2468-2667(19)30185-9.

10. Butler, H. J. et al. Using Raman spectroscopy to characterize biological materials. Nat. Protoc. 11, 664–687 (2016). https://doi.org/10.1038/nprot.2016.036

11. Ho, C. S. et al. Rapid identification of pathogenic bacteria using Raman spectroscopy and deep learning. Nat. Commun. 10, 4927 (2019). https://doi.org/10.1038/s41467-019-12898-9

12. Zuniga, W. C. et al. Raman Spectroscopy for Rapid Evaluation of Surgical Margins during Breast Cancer Lumpectomy. Sci. Rep. 9, 14639 (2019). https://doi.org/10.1038/s41598-019-51112-0

13. Desroches, J. et al. A new method using Raman spectroscopy for in vivo targeted brain cancer tissue biopsy. Sci. Rep 8, 1792 (2018). https://doi:10.1038/s41598-018-20233-3

14. Li, S. et al. Characterization and noninvasive diagnosis of bladder cancer with serum surface enhanced Raman spectroscopy and genetic algorithms. Sci Rep 5, 9582 (2015). https://doi.org/10.1038/srep09582

15. Sahu, A. K. et al. Oral cancer screening: serum Raman spectroscopic approach. J. Biomed. Opt. 20, 115006 (2015).

16. Jenkins, C. A. et al. A high-throughput serum Raman spectroscopy platform and methodology for colorectal cancer diagnostics. Analyst 143, 6014–6024 (2018). https://doi.org/10.1039/C8AN01323C

17. Kong, K., Kendall, C., Stone, N. & Notingher, I. Raman spectroscopy for medical diagnostics — From in-vitro bio fluid assays to in-vivo cancer detection. Adv. Drug Deliv. Rev. 89, 121–134 (2015).

18. Baker, M. J. et al. Clinical applications of infrared and Raman spectroscopy: state of play and future challenges. Analyst 143, 1735–1757 (2018). https://doi.org/10.1039/c7an01871a

19. Butler, H. J. et al. Development of high-throughput ATR-FTIR technology for rapid triage of brain cancer. Nat. Commun. 10, 4501 (2019). https://doi.org/10.1038/s41467-019-12527-5

20. NHS England, 2019 / 20 National Tariff Payment System. (2019). https://improvement.nhs.uk/resources/national-tariff/%h2-201920-national-tariff-payment-system (accessed 07/01/2020).

21. Coleman, M. P. et al. Cancer survival in Australia, Canada, Denmark, Norway, Sweden, and the UK, 1995–2007 (the International Cancer Benchmarking Partnership): An analysis of population-based cancer registry data. Lancet 377, 127–138 (2011).

22. Weller, D. et al. Diagnostic routes and time intervals for patients with colorectal cancer in 10 international jurisdictions; Findings from a cross-sectional study from the International Cancer Benchmarking Partnership (ICBP). BMJ Open 8, (2018).

23. National Institute for Health and Care Excellence. Quantitative faecal immunochemical tests to guide referral for colorectal cancer in primary care Diagnostics guidance. https://www.nice.org.uk/guidance/dg30 (2017). (accessed 01/12/19).

24. Quyn, A. J. et al. Application of NICE guideline NG12 to the initial assessment of patients with lower gastrointestinal symptoms: not FIT for purpose? Ann. Clin. Biochem. 55, 69–76 (2018).

25. Cohen, J. D. et al. Detection and localization of surgically resectable cancers with a multi-analyte blood test. Science 359, 926–930 (2018).

26. ACE Programme. Improving diagnostic pathways for patients with suspected colorectal cancer. (2017). https://www.cancerresearchuk.org/sites/default/files/ace-improvingdiagnosticpathwaysforpatientswithsuspectedcolorectalcancer-finalreportv1.0270617.pdf, (accessed 12/12/19).

27. Walter, F. M. et al. Evaluating diagnostic strategies for early detection of cancer: the CanTest framework. BMC Cancer 19, 1–11 (2019).

28. Harris, P. A. et al. Research electronic data capture (REDCap)— A metadata-driven methodology and workflow process for providing translational research informatics support. J. Biomed. Inform. 42, 377–381 (2009).

29. Tong, A., Sainsbury, P. & Craig, J. Consolidated criteria for reporting qualitative research (COREQ): A 32-item checklist for interviews and focus groups. Int. J. Qual. Heal. Care 19, 349–357 (2007).

30. Beleites, C., Neugebauer, U., Bocklitz, T., Krafft, C. & Popp, J. Sample size planning for classification models. Anal. Chim. Acta 760, 25–33 (2013).

31. Thorne, K., Hutchings, H. A. & Elwyn, G. The effects of the Two-Week Rule on NHS colorectal cancer diagnostic services: A systematic literature review. BMC Health Serv. Res. 6, 1–5 (2006).

32. Buderer, N. M. F. Statistical Methodology: I. Incorporating the Prevalence of Disease into the Sample Size Calculation for Sensitivity and Specificity. Acad. Emerg. Med. 3, 895–900 (1996).

33. Malhotra, R. K. & Indrayan, A. A simple nomogram for sample size for estimating sensitivity and specificity of medical tests. Indian J. Ophthalmol. 58, 519–522 (2010).

34. R Core Team. R: A language and environment for statistical computing. R Foundation for Statistical Computing, Vienna, Austria. http://www.r-project.org/index.html. (2019).

35. Breiman, L. Random forests. Mach. Learn. 5–32 (2001). https://doi.org/10.1201/9780429469275-8

36. Fidler, J. L. et al. Abdominal Imaging Detection of flat lesions in the colon with CT colonography. Abd. Imag. 300, 292–300 (2002).

37. Macari, M. et al. Radiology Colorectal Neoplasms: Prospective Comparison of Thin-Section Low-Dose Multi - Detector Row CT Colonography and Conventional Colonoscopy for Detection. Radiology 2, 383–392 (2002).

38. Smith, J. & Firth, J. Qualitative data analysis: the framework approach. Nurse Res. 19, 52–62 (2011).

